# Comparative modeling of fetal exposure to maternal long-acting injectable versus oral daily antipsychotics

**DOI:** 10.1101/2024.10.22.24315924

**Authors:** Philip Bediako-Kakari, Mariella Monyo, Shakir Atoyebi, Adeniyi Olagunju

**Author notes:** **Corresponding author:** Dr Shakir Atoyebi. Department of Biochemistry, Cell, and Systems Biology, Biosciences Building, Crown Street, Liverpool, L69 7BE, United Kingdom. Both PB and MM are joint first-authors.

## Abstract

This study employed physiologically based pharmacokinetic (PBPK) modelling to compare the extent of fetal exposure between oral and long-acting injectable (LAI) aripiprazole and olanzapine. Adult and pregnancy PBPK models were developed and validated with relevant clinical data. Relevant indices of fetal exposure during pregnancy were predicted from concentration-time data at steady-state dosing for both oral and LAI formulations. Fetal C_max_ for aripiprazole was 59-78% higher with LAI than oral, and 68-181% higher with LAI olanzapine than the oral formulation. Predicted C:M ratios (range) was 0.59-0.69 for oral aripiprazole and 0.61-0.66 for LAI aripiprazole, 0.34-0.64 for oral olanzapine and 0.89-0.96 for LAI olanzapine. Also, cumulative fetal exposure over 28 days from oral formulations were generally predicted to be lower compared with their therapeutic-equivalent LAI. As *in utero* fetal exposure to maternal drugs does not necessarily translate to risk, these data should be interpreted in a broader context that includes benefit-risk assessments.

## Introduction

Around 1.5 to 3.5% of the general population will be diagnosed with a psychotic disorder, while a higher percentage will experience at least one psychotic symptom at some point in their lives^1^. Implications of poor treatment of psychosis leads to significant repercussions not only for the affected individuals and society. Such implications could include worsening of symptoms, functional decline^2^, increased risk of suicide^3^, physical health deterioration^4^, increased hospitalizations^2^, social isolation and stigma^5^, burden on families, caregivers and healthcare services as well as legal and forensic issues^6,7^.

Oral antipsychotic drugs remain the standard treatment option for psychosis. These drugs, classified into first-generation or second-generation antipsychotics, function by altering neurotransmitter activity in the brain to reduce symptoms such as hallucinations and delusions. While oral antipsychotics are widely used, their effectiveness can be underutilized by poor adherence to medication regimens, particularly in individuals with severe mental illness who may have cognitive deficits or lack insight into their condition^8-10^. Additionally, oral medications are subject to first-pass metabolism in the liver, leading to variable bioavailability and a significant portion of the drug being metabolized before reaching systemic circulation^11,12^. This variability may necessitate higher doses to achieve therapeutic effects^13,14^, increasing the risk of side effects^15,16^.

Long-acting injectables (LAIs) were developed to address the limitations associated with oral antipsychotics, particularly adherence and bioavailability issues, by offering consistent drug delivery system^17^. They ensure guaranteed administration and transparency of adherence by eliminating the need for daily dosing^18,19^. These lead to improved treatment outcomes, as LAIs also reduce the likelihood of missed doses, unintentional overdose, and abrupt relapses due to partial or overt nonadherence^20-22^. Additionally, by bypassing gastrointestinal absorption, LAIs avoid first-pass metabolism ^21,22^ leading to consistency in bioavailability^23^, resulting in reduced peak-trough plasma levels^17^,^24^. However, LAIs also present challenges, such as slow dose titration, injection-site discomfort, and frequent visits for treatment^17^. These issues may be amplified during pregnancy, where physiological changes can alter drug disposition^25-27^.

Pregnancy presents a period of significant psychological ^28,29^ and physiological change^26,27^, and for women with pre-existing mental health conditions, an increased risk of exacerbation or relapse^25. 25,30,31^As such, managing psychosis during pregnancy is crucial, as untreated mental illness can harm both the mother and the fetus, leading to poor prenatal care, substance abuse, and obstetric complications^32^. Pregnancy can further complicate oral medication use due to nausea, vomiting, and difficulty swallowing, particularly during the first trimester^33^. Additionally, there is a risk of inadvertent fetal exposure to antipsychotics^34^, which can be associated with low birth weight, preterm birth, and congenital malformations^35,36^. Despite these risks, continuing antipsychotics is often necessary for maternal and fetal health^36^. However, there is limited knowledge on how fetal exposure differs between oral and long-acting injectable (LAI) antipsychotics and understanding these differences could help reduce the risk of congenital issues.

Estimating fetal exposure is challenging due to the ethical and practical limitations on direct sampling of fetal blood. Consequently, various indirect methods have been used to predict fetal drug levels including pharmacokinetic modelling, placental tissue sampling, cord blood sampling at delivery, and the use of animal models. Among these, physiologically based pharmacokinetic (PBPK) models have gained prominence, allowing for the simulation of *in utero* maternal-to-fetal drug transfer throughout pregnancy. These models integrate physiological changes occurring in both the mother and the fetus during gestation^37,38^, providing a dynamic approach to estimate fetal drug exposure across different stages of pregnancy^39^.

Aripiprazole and olanzapine are commonly prescribed antipsychotics, targeting serotonin (5-HT) and dopamine receptors. Aripiprazole is metabolized by CYP2D6 and CYP3A4 into its active metabolite dehydro-aripiprazole, while olanzapine is metabolized mainly through CYP1A2, CYP2C8, and UGT1A4. Both drugs are available in oral once-daily tablets (aripiprazole: 2 – 30 mg; olanzapine: 2.5-20 mg) and LAI formulations. The LAI formulation of aripiprazole is available in two distinct forms: aripiprazole once-monthly (AOM) and aripiprazole lauroxil. AOM is a monohydrate version, while aripiprazole lauroxil is a prodrug with a higher molecular weight (660.7 g/mol versus 466.4 g/mol)^40,41^. AOM comes in 300 mg and 400 mg doses, equivalent to 15 mg and 20 mg daily doses, respectively^42,43^.^41^ The LAI formulation of olanzapine contains olanzapine esterified with pamoic acid which confers sustained-release properties enabling four-weekly intramuscularly (IM) administration at a dose of 300mg or 405mg^44^.

In this study, we used PBPK modelling to compare fetal exposure during pregnancy between oral antipsychotics and their therapeutic-equivalent LAI formulations.

## Methodology

### Model description

Our previous whole-body adult and pregnancy PBPK models developed for long-acting cabotegravir and rilpivirine, were repurposed and used for this study ^57^. Both models were implemented in SimBiology®, an application within MATLAB® (version R2023b, MathWorks, Natick, USA, 2023). The pregnancy PBPK model had been extrapolated from the whole-body adult model by incorporating the various gestation-related anatomical and physiological changes, as well as pregnancy-specific compartments.

### System parameters

Anatomical and biological parameters such as organ volumes, blood flow rates, and tissue composition used in the creation of the model were obtained from available literature. The weights of the various organs and tissues were determined via anthropometric equations previously reported by Bosgra et al. (2012) and these were combined with reported organ densities (Brown et al., 1997) to estimate organ volumes. Blood flow rates to and from the various tissues and organs were also parameterised as fractions of the cardiac output as previously reported in literature^45^.

### Absorption

Oral drug absorption was modelled with the compartmental absorption and transit model^46^. The oral absorption rate constant (K_A_) was estimated using the effective drug permeability (***Equation 1***), which was derived from the number of hydrogen bond donors (HBD) and the polar surface area (PSA) of the aripiprazole and illustrated in ***Equation 2***^***47***^. For olanzapine, K_A_ was obtained from literature^48^.

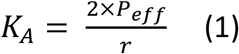

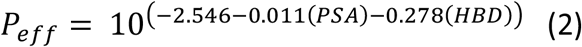

where r represents the radius of small intestines.

The gut abundance ^49^ and intrinsic clearance of CYP3A4^50^, were used to calculate the gut clearance (Cl_gut_) of drug. The fraction of drug escaping intestinal metabolism and reaching the liver was modelled using ***Equation 3***.

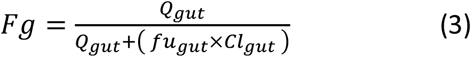

where fu_,gut_ is the fraction of unbound drug and is modelled as 1, and Q_gut_ (L/h) denotes the gut blood flow.

The intramuscular release of aripiprazole LAI formulation from the depot compartment in the muscle was simulated as a first-order reaction, described by ***Equation 4***.

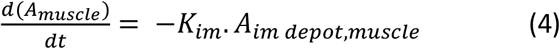

where d(A_muscle_)/dt, represents the aripiprazole release rate constant from the IM depot into the systemic circulation (mg/h), K_im_ denotes the IM aripiprazole rate release constant from IM depot, and A_im depot,muscle_ indicates the quantity of aripiprazole (mg) present in the IM depot within the muscle.

However, for olanzapine the intramuscular LAI release rate better described using ***Equation 5***. The two different rates were rationalised as representing immediate release into circulation post-administration and a long-term gradual release from intramuscular depot.

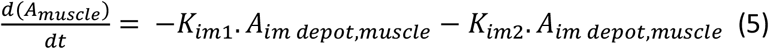

where d(A_muscle_)/dt, represents the olanzapine release rate constant from the IM depot into the systemic circulation (mg/h), A_im depot,muscle_ indicates the quantity of olanzapine (mg) present in the IM depot within the muscle.

### Distribution

The schematic diagram of the pregnancy PBPK model is illustrated in **Figure 1(A)** showing the fetal component within the uterus. Key model assumptions included perfusion-limited drug distribution and the well-stirred distribution^39^. Volume of drug distribution (V_ss_) was modelled using the tissue-to-plasma ratios and volumes of the various compartments^51^. The impact of pregnancy on the fraction of unbound drug was modelled using equations from existing literature^52,53^.

**Figure 1.**
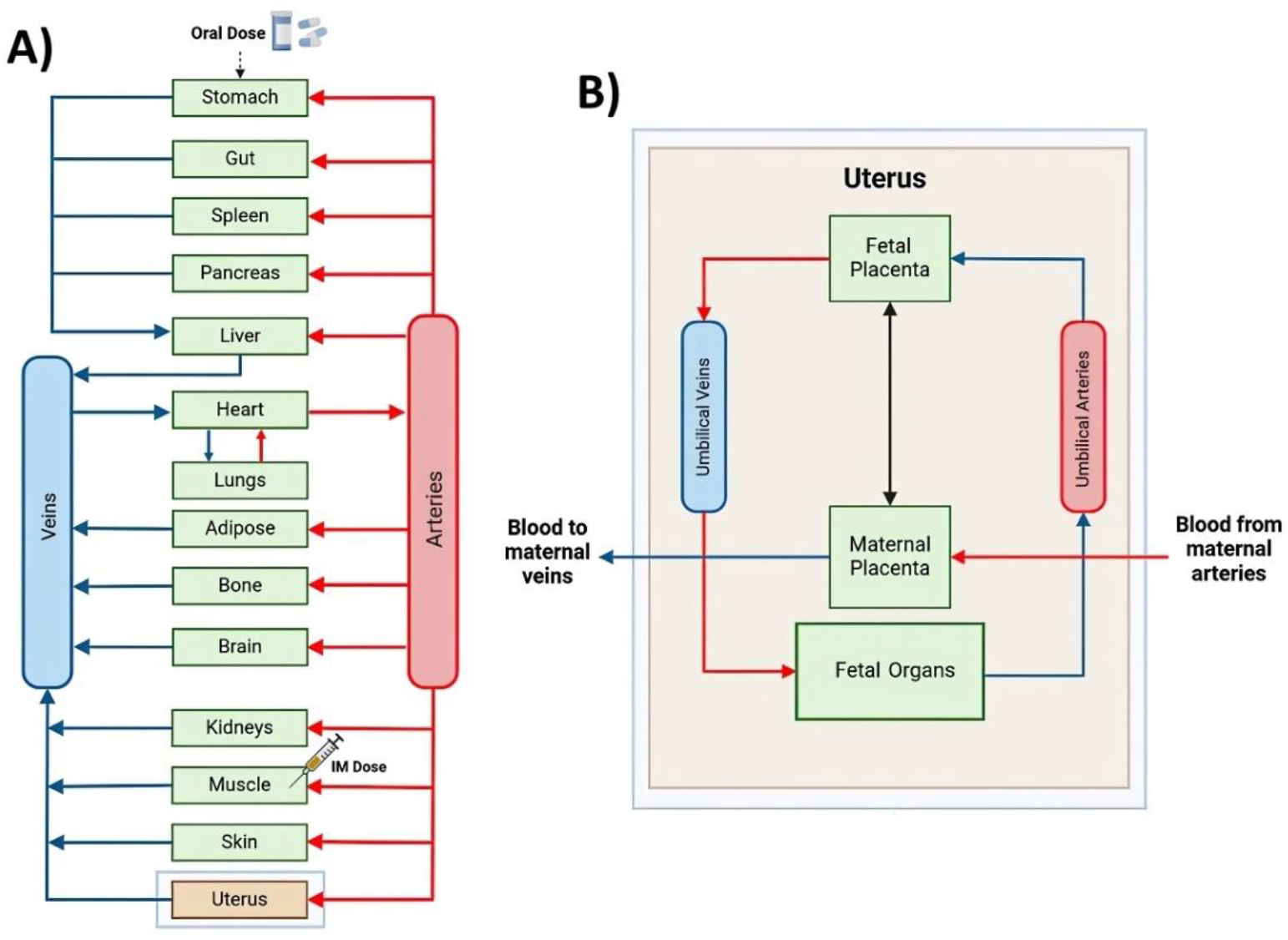
(A) Illustration of the physiologically-based pharmacokinetic (PBPK) model representing tissues and organs as compartments with the arrows indicating the direction of blow flow. (B) Schematic diagram of the fetal compartment as modelled in the uterus. (Adapted from Atoyebi, et al. ^39^ and recreated on Biorender.com).

### Metabolism

Aripiprazole metabolism was modelled as previously reported in literature^50^. Based on in-vitro studies, aripiprazole undergoes three biotransformation pathways mediated by CYP2D6 and CYP3A4: hydroxylation, N-dealkylation and dehydrogenation^54^. Therefore, the total liver clearance was modelled using reported drug clearances by each enzyme^50^. Thus, the liver clearance was modelled using ***Equation 6***.

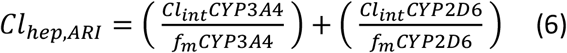

where Cl_int_CYP3A4 is the intrinsic clearance by the CYP3A4 enzyme; Cl_int_CYP2D6 is the intrinsic clearance by the CYP2D6 enzyme; Cl_hep,ARI_ is the total liver clearance of aripiprazole, f_m_CYP3A4 is the fraction metabolized by CYP3A4 and f_m_CYP2D6 is the fraction metabolized by CYP2D6. For olanzapine metabolism, liver clearance was determined based on the intrinsic clearance of the drug by the following enzymes: CYP1A2, CYP2C8, CYP3A4, FMO3, and UGT1A4 (81). The intrinsic clearance of the drug per milligram of microsomal protein for each enzyme was scaled to the entire liver, accounting for the microsomal protein content as well as total liver weight (***Equation 7-8***).

However, the renal olanzapine clearance was not included in the model.

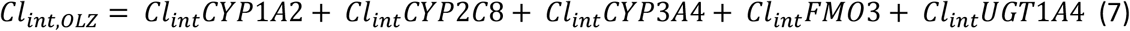

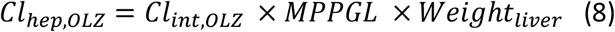

Where Cl_int,OLZ_ is the total intrinsic clearance of olanzapine; Cl_int_CYP1A2 is the intrinsic clearance by the CYP1A2 enzyme; Cl_int_CYP2C8 is the intrinsic clearance by the CYP2C8 enzyme; Cl_int_CYP3A4 is the intrinsic clearance by the CYP3A4 enzyme; Cl_int_FMO3 is the intrinsic clearance by the FMO3 enzyme; Cl_int_UGT1A4 is the intrinsic clearance by the UGT1A4 enzyme; Cl_hep,OLZ_ is the total liver clearance of olanzapine; MPPGL is the weight of microsomal protein per gram of liver and, Weight_liver_ is the total weight of the liver (kg).

Fraction of drug escaping the liver into the systematic circulation (Fh) was determined by using the total liver clearance (Cl_hep_), as described in ***Equation 9***.

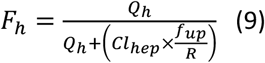

where Q_h_ denotes hepatic blood flow rate, f_up_ represents the fraction of unbound aripiprazole and olanzapine in plasma and R indicates the tissue-to-plasma ratio of drug in the liver.

### Pregnancy PBPK model

The whole-body pregnancy PBPK model was similar to the pregnancy PBPK model reported by Atoyebi, et al. ^39^. The development of the pregnancy PBPK model included involved feminising the adult PBPK model by restricting gender-specific parameters to reflect female-only characteristics. Additionally, the model was modified to incorporate pregnancy-induced anatomical and physiological changes known to influence drug disposition^55,56^. The rate of blood flow to various tissues and organs were calculated as fractions of cardiac output, using data from available literature^56^. Furthermore, gestation-related changes to the metabolic activity of CYP3A4 and CYP2D6 as reported by Abduljalil, et al. ^55^, were implemented into the model, ***Equations (10-11)***.

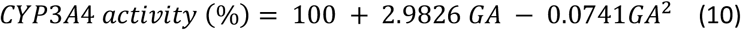

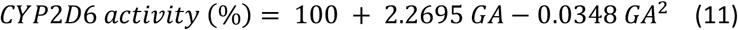

Where GA denotes gestational age.

### Drug Parameters

The PBPK models were refined to simulate both aripiprazole and olanzapine metabolism by incorporating key drug-specific parameters reported in literature. These included the acid dissociation constant, the octanol-water partition coefficient, and the first-order release rate constant for the intramuscular depot, which play significant roles in simulating drug absorption and distribution. Presented in Table S1 is an overview of the parameters, including both the in vitro parameters and physico-chemical properties of aripiprazole and olanzapine implemented in the models.

### Validation of PBPK Models with Clinical Data

For each scenario, simulations were conducted in 100 virtually healthy population with dosages, routes of administrations, and demographics, set as reported in the clinical studies used for model validation. Predicted drug concentration-time data were analysed for dosing intervals of interest for each simulation and used to determine pharmacokinetic (PK) parameters, including the area under the curve (AUC_t_), maximum concentration (C_max_), minimum concentration (C_min_), and half-life (T_1/2_). Predicted PK parameters were compared against observed data reported in the clinical studies. A prediction error within a 2-fold range was considered the acceptable limit for this study, accounting for the any variability in predicted PK parameters, as outlined by Abduljalil, et al. ^57^.

### Aripiprazole

For validation of the whole adult PBPK model for aripiprazole, clinical data from non-pregnant populations receiving oral and LAI aripiprazole were used. For the oral formulation, clinical PK data from 5 mg and 10 mg single oral dose of aripiprazole were used, as reported by Boulton, et al. ^58^ in 14 participants over 384 hours and by Wojnicz, et al. ^59^ in 103 healthy volunteers over 72 hours, respectively. Similarly, clinical PK data from Raoufinia, et al. ^42^ were used for the validation of LAI aripiprazole, specifically focusing on single and repeated monthly administrations of 400 mg AOM administered via both gluteal and deltoid injections in patients with schizophrenia. In the single administration study, both cohorts receiving the 400 mg aripiprazole dose via deltoid (n = 18) and gluteal (n = 19) administration were monitored for over 126 days. For the repeat-administration study, participants were observed for 141 days, with two different administration sequences: deltoid/deltoid (n = 73) and gluteal/deltoid (n = 68). All steady-state PK samplings in the repeat-dose study were conducted after the fifth injection.

Due to the limited availability of clinical data in pregnant populations, no literature specifically reported the PK of aripiprazole in either oral or LAI, in this group. Instead, timepoint concentrations for each trimester, as reported by Westin et al. (2018) on various antipsychotics, including aripiprazole, in pregnant women, were used to validate the pregnant model. The participants (n = 14) were on 15 mg once daily aripiprazole until the end of term and had reached steady-state concentrations at the time of PK sampling.

### Olanzapine

The adult model was validated with clinical PK data for olanzapine reported by Du et al. (2020), where a single dose of 5mg was orally administered to healthy volunteers under fed (n = 24) and fasting (n = 30) conditions. For validation of single dose of 10 mg, clinical PK data reported by Sun et al. (2019) included healthy male and female volunteers (n = 45) observed for a period of 168 hours (Sun et al., 2019). For LAI, clinical data from 300 mg bi-weekly injections (n = 19) and 405 mg monthly injections (n = 29) of olanzapine in patients with schizophrenia over a period of 24 weeks as reported by Mitchell et al. (2013).

For the validation of the pregnancy PBPK model for olanzapine, clinical data used were drug plasma concentration-time data reported by Westin, et al. ^60^ for orally administered 10 mg olanzapine in patients with psychosis (n = 29) across all three trimesters during pregnancy.

### Fetal Compartment Modelling

A multi-compartment fetal sub-model was integrated into the uterus compartment to create the materno-fetal PBPK (m-f-PBPK) model. It includes compartments representing maternal-placenta, fetal-placenta, fetal liver, rest of the fetus, umbilical veins and umbilical arteries (Figure 1B).

Drug movement within the fetal sub-model was implemented as following a defined path where drugs within the maternal bloodstream were delivered to the maternal side of the placenta via the maternal arteries. At the placenta, drugs cross the placental barrier and exchange with the fetal blood. This exchange via passive diffusion allows drug transfer to the fetal side of the placenta. From there, the drugs are carried through the umbilical vein towards the fetal organs, passing through the fetal ductus venosus and the fetal portal sinus, where they are distributed to the fetal tissues. After circulating through the fetal organs, the remaining drugs in the blood, is collected via the umbilical arteries and carried back to the placenta. At the placenta, another exchange occurs across the placental membrane, allowing the available drugs to transfer to the maternal side via passive diffusion. Finally, these substances transferred back to the maternal veins for excretion and or further metabolism by the maternal body.

Blood flow to fetal organs was modelled using equations described by Zhang, et al. ^38^. The equation defining blood flow through the portal sinus was parameterised as the difference between the blood flow in the ductus venosus and the umbilical vein. The blood flow through the fetal hepatic vein was modelled as the total blood flow through fetal hepatic vein and fetal portal vein. Similarly, the blood flow through the fetal liver was characterised by the summation of the blood flow through the fetal hepatic vein and fetal portal sinus. Presented in **Table 1** are equations of the various gestation-related changes to blood flows in the various blood vessels, as well as various fetal organs implemented in the fetal model^38,55^.

**Table 1:**
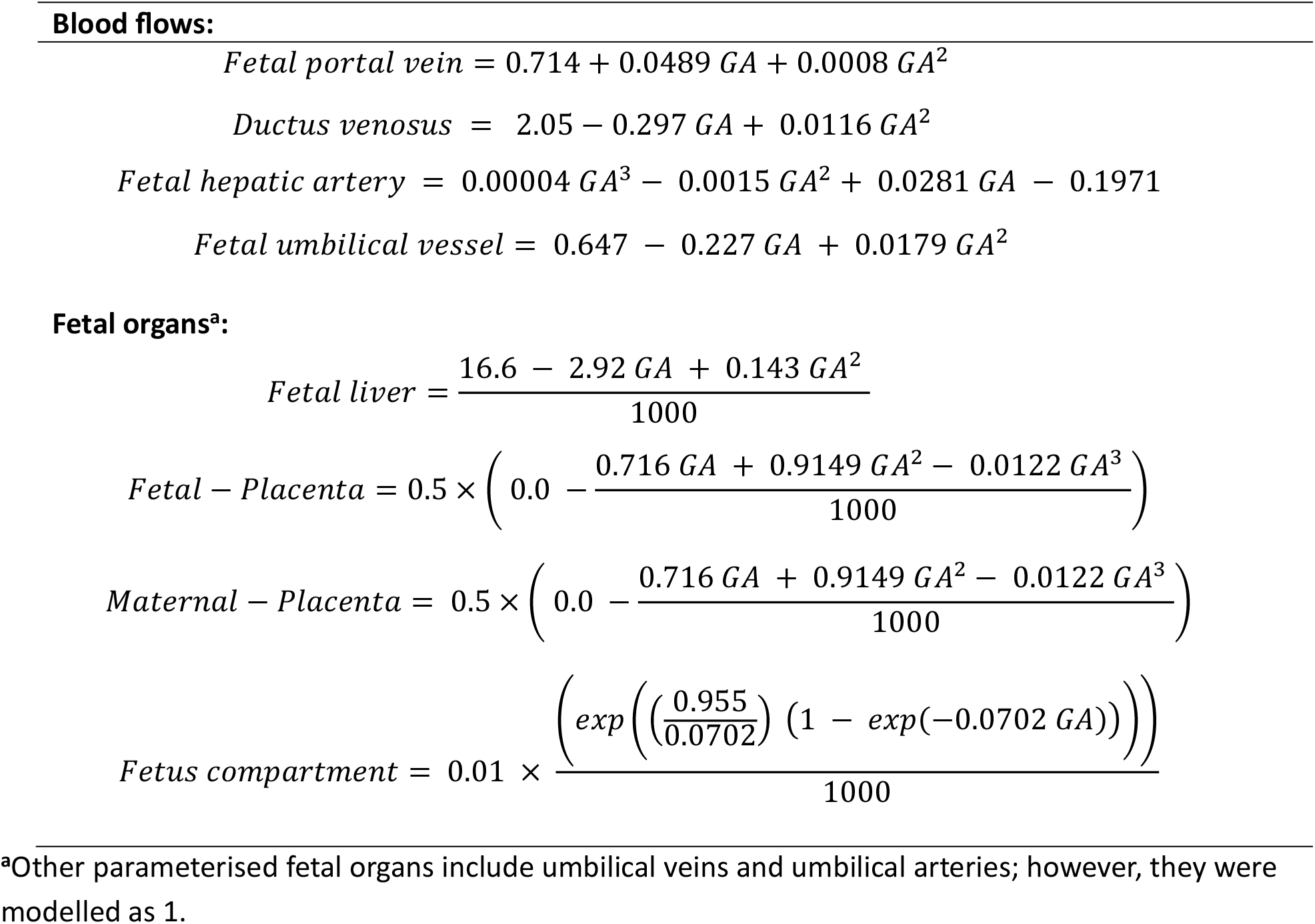
Equations for blood flows and organ measurements used in the fetal model.

The bidirectional passive diffusion of the drugs (Q_pd_) across the placenta was established as described by Zhang, et al. ^38^. The diffusion rate across the placenta ***Equation 12*** was quantified by adapting Fick’s law of diffusion^38,61^.

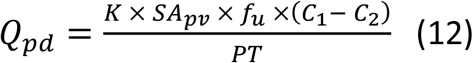

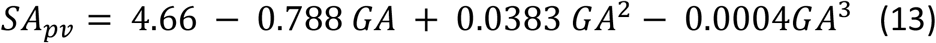

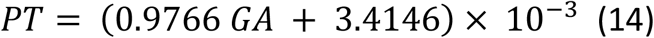

where K denotes the diffusion rate constant, which was fitted to 17 for aripiprazole and 0.0108 for olanzapine. SA_pv_ represents the surface area of the placental villous (m^2^), f_u_ is the fraction of the unbound, (C_1_ - C_2_) denotes the transplacental concentration gradient, PT represents the thickness of the placenta (m), and GA represents gestational age (weeks).

Fetal liver clearance was parameterised by including CYP enzymes present in the fetal liver using ***Equation 14***. The fetal intrinsic clearance of CYP3A4 was extrapolated from the adult intrinsic clearance values. Additionally, the fetal abundance of CYP3A4, mass per gram of fetal liver, and fetal liver weight were all sourced from available literature^62-64^.

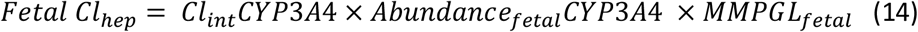

Where Cl_int_CYP3A4 is the adult intrinsic clearance by the CYP3A4 enzyme, Abundance_fetal_CYP3A4 is the fetal abundance of CYP3A4, and MMPGL_fetal_ is the mass per gram of fetal liver (mg/g).

### Validation of the m-f-PBPK model

Using the m-f-PBPK model, simulations were performed to validate the model predictions of drug transfer to the fetal compartment. For aripiprazole, these simulations were compared against a case study reported by Nguyen et al. (2011), involving a 27-year-old mother with schizophrenia who was administered 10 mg of the drug at 37 weeks gestation and was considered to have reached steady-state at delivery (39.3 weeks), the time of PK sampling. To validate the maternal-fetal dyad for olanzapine, a single case study reporting fetal and maternal concentration at delivery was also used^65^. Given that the pregnancy model was able to adequately predict fetal exposure to oral aripiprazole and olanzapine in the pregnant population across all three trimesters and at delivery, it was assumed that model predictions for fetal exposure to LAI drugs would also be adequate.

### Modelling fetal exposure to oral versus LAI formulations

Reported oral aripiprazole dosages of 15 mg and 20 mg have shown PK profiles comparable to those of LAI aripiprazole at 300 mg and 400 mg, respectively^42,43^. Additionally, the most recommended starting oral dose for olanzapine is 10 - 15 mg once daily, with corresponding LAI doses being 300 and 405 mg once monthly respectively^66-68^. Thus, simulations were conducted to predict the pharmacokinetics of the oral dosages (e.g., 15 mg ARI and 15 mg OLZ) and their therapeutic-equivalent LAI doses (300 mg ARI and 405 mg OLZ, respectively) in virtual pregnant populations (n = 100 per scenario). The mean (SD) body weight and age of the virtual population were 57.9 (12.6) kg and 31.5 (7.9) years, respectively.

For oral doses, one dosing interval (within 24 hours) at steady state was analysed during the second and third trimesters. Similarly, steady-state pharmacokinetics of LAI aripiprazole were simulated during the second and third trimesters with analysis carried-out for a monthly dosing interval. Fetal exposure was estimated by calculating the time-averaged cord plasma concentration relative to maternal plasma concentration ratio (C:M ratio). Also, the fetal-to-maternal exposure ratio within each dosing interval was estimated by dividing the area under the drug concentration curve (AUC) of the fetus by that of the mother (fAUC/mAUC). Variations in fetal and maternal plasma concentrations over one dosing interval at steady state were also assessed.

## Results

### Adult models validation

The observed and simulated PK parameters for both orally and intramuscularly administered aripiprazole and olanzapine were compared, as shown in **Table S2**. The PBPK model was successfully validated for both aripiprazole and olanzapine PK, with the calculated absolute average fold error (AAFE) for all parameters within the set 2-fold threshold. Likewise, the pregnancy PBPK model adequately predicted both aripiprazole and olanzapine timepoint concentrations at each trimester (**Table S3**).

### Pregnancy models validation

Simulations performed to estimate fetal drug exposure successfully predicted cord plasma and maternal plasma concentrations for both aripiprazole and olanzapine within the set acceptable range, as shown in **Table S4**. All calculated AAFEs were below the 2-fold threshold, confirming the adequacy of the PBPK model to simulate fetal exposure to both aripiprazole and olanzapine after maternal dose during pregnancy.

### Fetal exposure simulations

Predicted indices of fetal exposure at steady state summarized as C:M ratio and other predicted PK parameters of aripiprazole and olanzapine are shown in **Table 2**. Generally, fetal exposure was predicted to decrease with increasing gestation in pregnancy for both drugs.

**Table 2:**
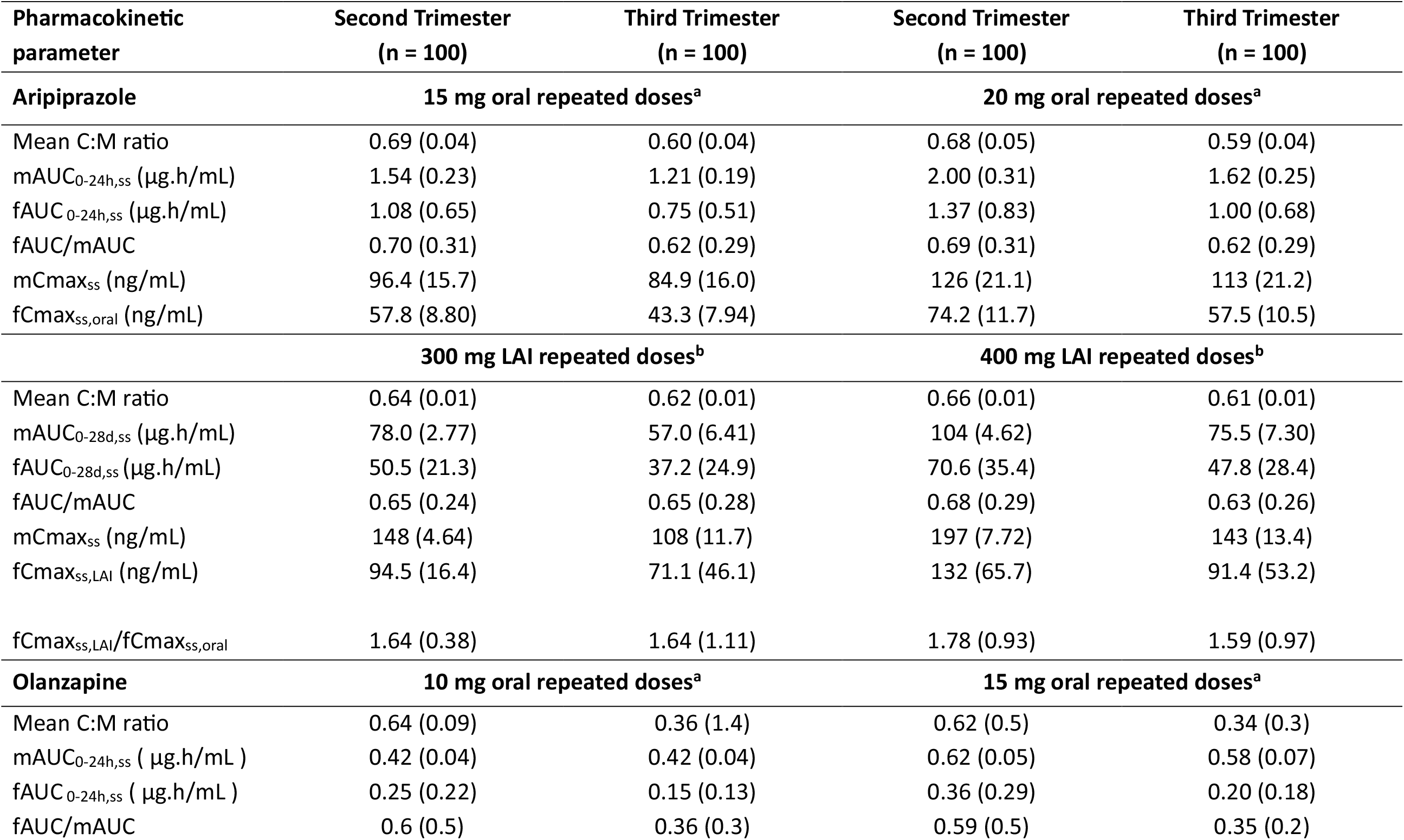
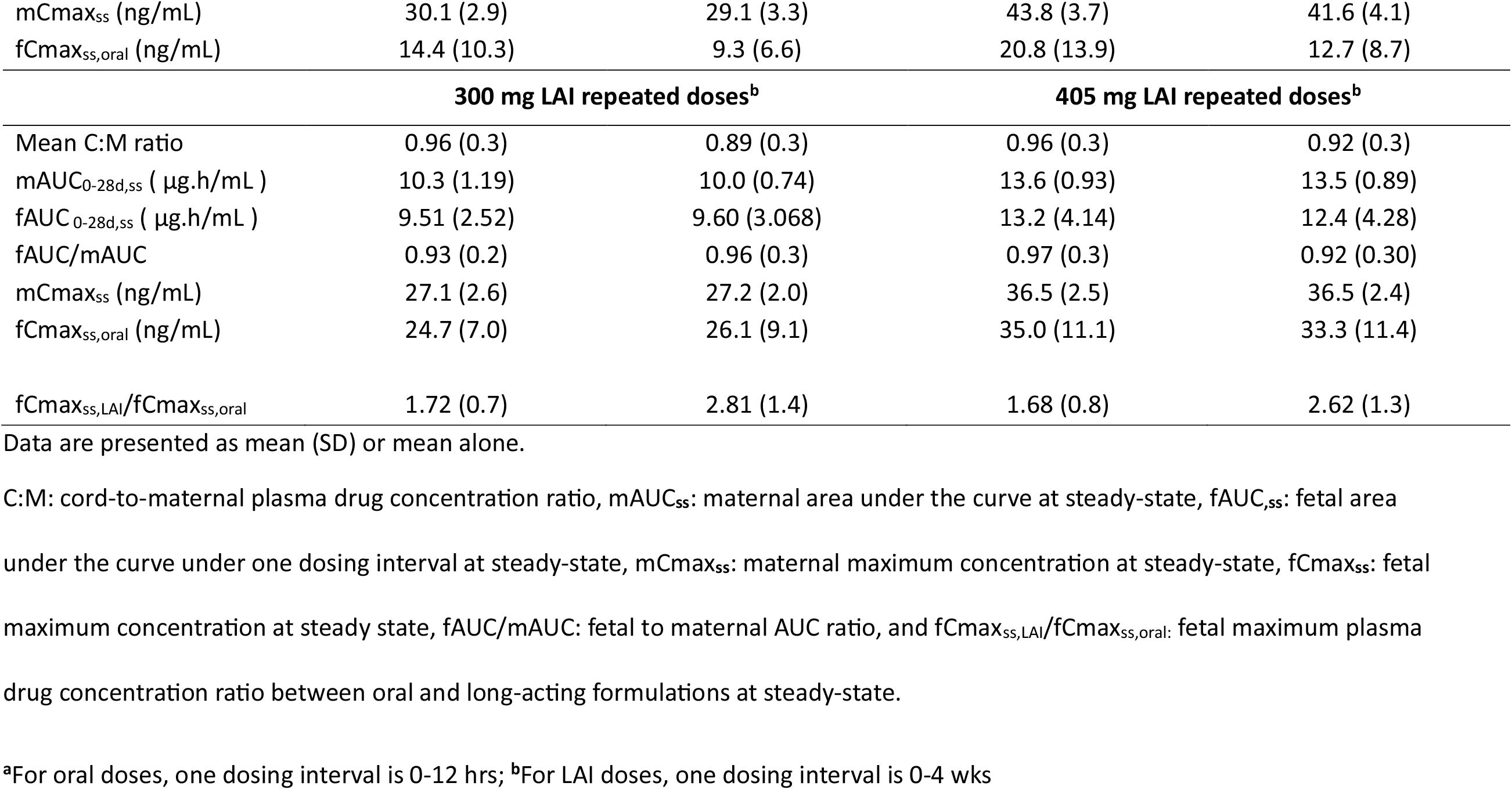
Predicted pharmacokinetics parameters of repeated oral versus therapeutic-equivalent LAI aripiprazole doses at steady state during pregnancy.

Predicted C:M ratios for oral and LAI aripiprazole and olanzapine are shown in **Figure S1 Figure S1** and **Figure S2**, respectively. Predicted C:M ratios at steady state within a dosing interval decreased with gestational age. For the oral doses, predicted C:M ratios also fluctuated within each dosing interval, rising shortly after each new dose and peaking at approximately 4hrs post-dose for aripiprazole and 5hrs post-dose for olanzapine, after which they plateaued until the end of the dosing interval. Similar to the oral drugs, predicted C:M ratios for both LAI drugs at steady state also decreased with gestational age, but with less fluctuations within the dosing intervals. Across trimesters and formulations, the predicted C:M ratios for both drugs were consistently below 1. Predicted median (range) C:M ratio was 0.63 (0.59-0.69) for aripiprazole and 0.77 (0.34-0.96) for olanzapine.

Changes in predicted maternal and fetal plasma concentrations for aripiprazole and olanzapine in a single dosing interval are shown in **Figure 2 and Figure 3**. The figures illustrate predicted fetal plasma concentrations are consistently lower than maternal plasma concentrations for both drugs. Furthermore, these drug plasma concentrations decrease as pregnancy progresses. The predicted mean fetal AUC for 10 mg oral olanzapine in second trimester was 0.25 (increasing to 7 μg.h/mL when extrapolated to 28 days), and for 20 mg oral aripiprazole it was 1.37 (increasing to 38.36 μg.h/mL when extrapolated to 28 days).

**Figure 2.**
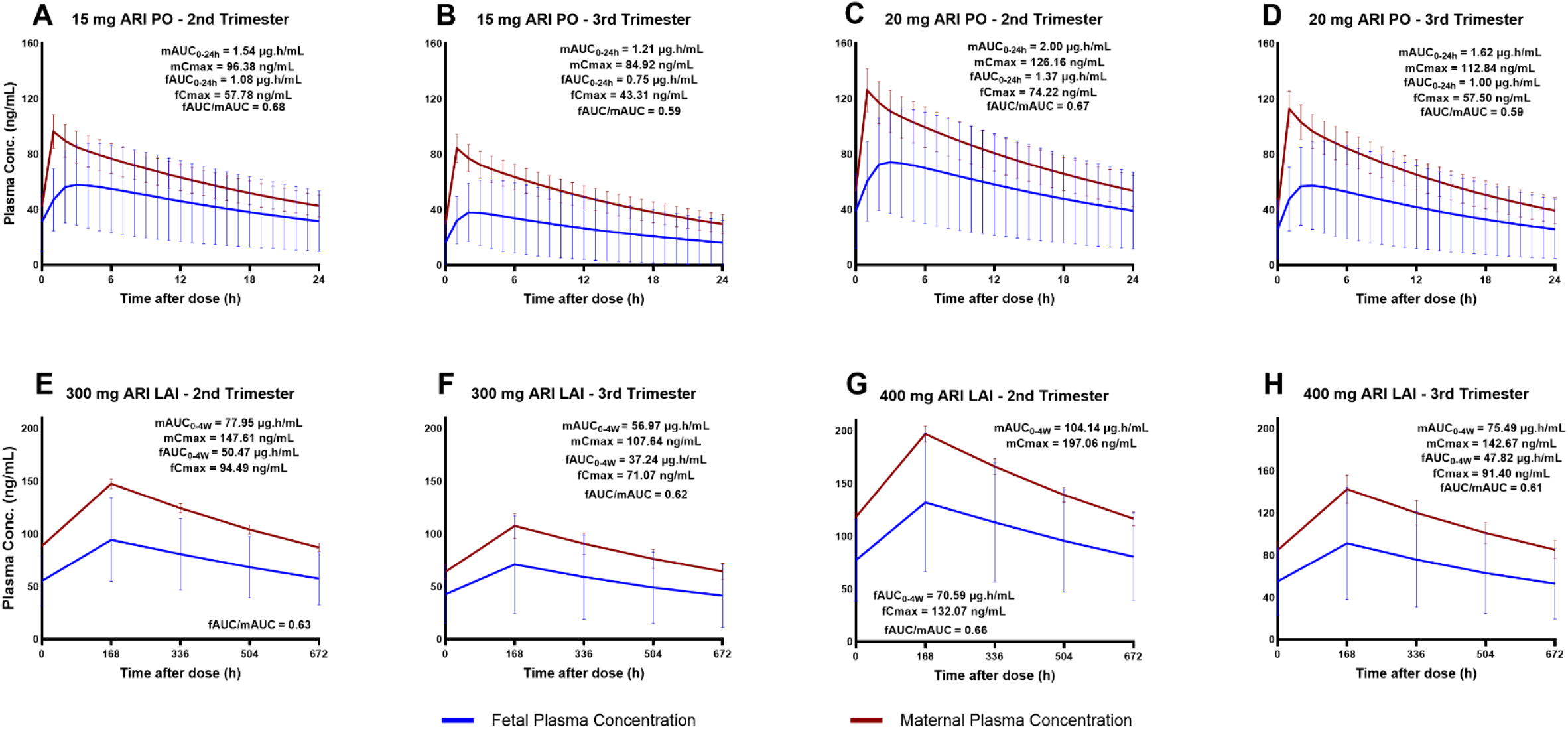
Predicted average plasma concentration-time profile of (PO) and their therapeutic-equivalent long-acting injectable (LAI) doses of aripiprazole in pregnancy in one dosing interval at steady-state. **(A)** 15 mg oral dose in the second trimester, **(B)** 15 mg oral dose in the third trimester, **(C)** 20 mg oral dose in the second trimester, **(D)** 20 mg oral dose in the third trimester **(E)** 300 mg LAI dose in the second trimester, **(F)** 300 mg LAI dose in the third trimester, **(G)** 400 mg LAI dose in the second trimester, **(H)** 400 mg LAI in the third trimester.

**Figure 3.**
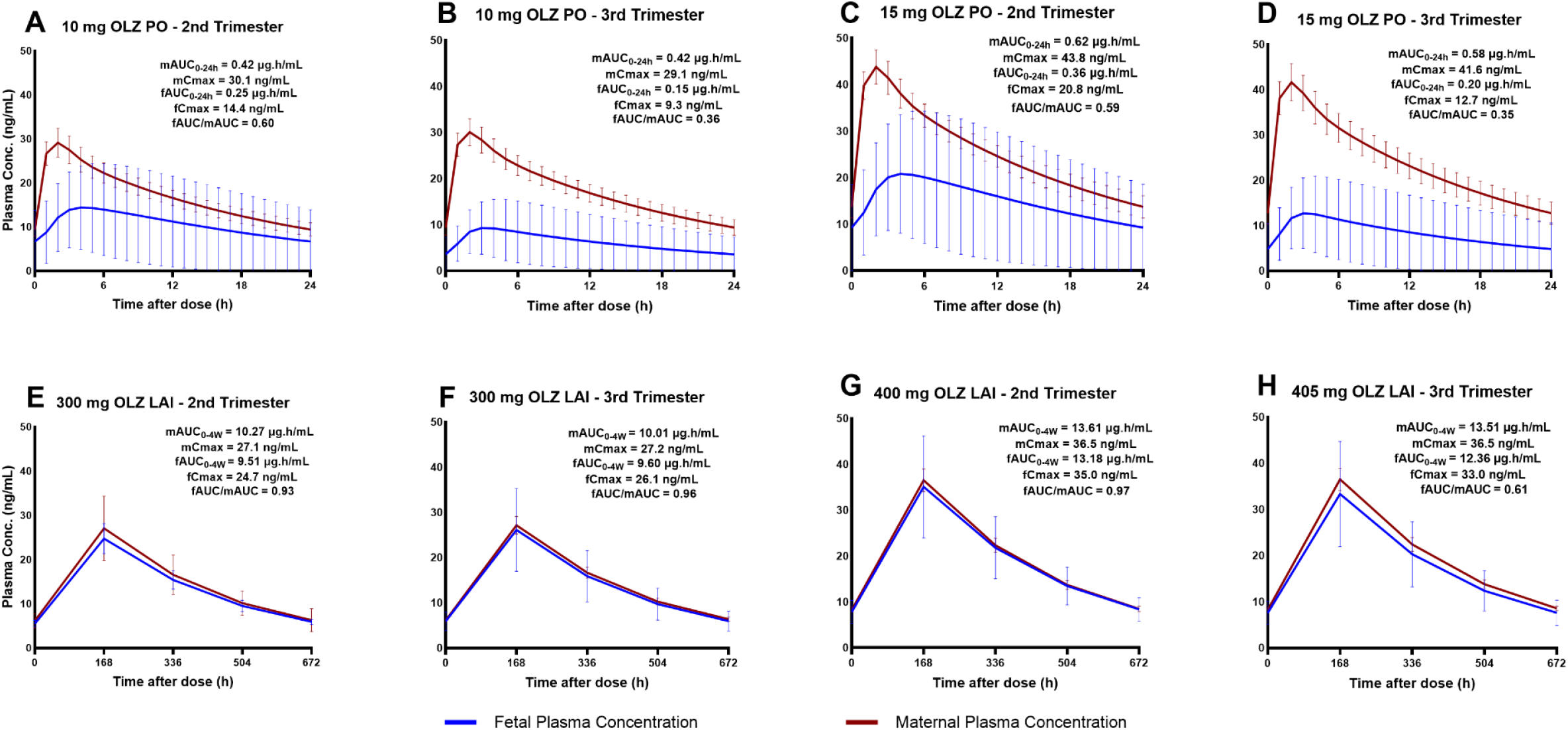
Predicted average plasma concentration-time profile of (PO) and their therapeutic-equivalent long-acting injectable (LAI) doses of olanzapine in pregnancy in one dosing interval at steady-state. **(A)** 10 mg oral dose in the second trimester, **(B)** 10 mg oral dose in the third trimester, **(C)** 15 mg oral dose in the second trimester, **(D)** 15 mg oral dose in the third trimester, **(E)** 300 mg LAI dose in the second trimester, **(F)** 300 mg LAI dose in the third trimester, **(G)** 405 mg LAI dose in the second trimester, **(H)** 405 mg LAI in the third trimester.

## Discussion

In this study, the extent of in utero exposure was compared between the respective oral and long-acting injectable (LAI) formulations. So far, only one PBPK model has been utilized to predict drug disposition of both aripiprazole^50^ and olanzapine^68^ in pregnant populations. To the best of our knowledge, this is the first study to compare in utero exposure between oral and LAI formulations of both aripiprazole and olanzapine through PBPK modelling.

Comparative analysis of oral and LAI formulations of both aripiprazole and olanzapine did not show any notable difference across trimesters based on the C:M and fAUC/mAUC ratios (**Table 2**). The simulated C:M ratios consistently remained below 1 for both drugs This suggests that irrespective of dose or formulation the portion of the drug crossing the placenta to the fetus is lower compared to the amount of the drug in the mother.

Simulated C:M ratios for oral and LAI formulations of aripiprazole, remained relatively consistent. However, this consistency was not observed with olanzapine, as both the C:M and fAUC/mAUC ratios varied between the two formulations. A study by Correll, et al. ^69^ reported the peak-to-trough plasma concentration of oral olanzapine in comparison to its LAI once monthly formulation has an approximate 2-fold difference accounting for the disparities in PK profiles.

While C:M ratios might offer insight into the extent of fetal exposure at a specific time, it is inadequate to quantify the total amount of fetal drug exposure over time. Predicted mean C_max_ decreased with gestational age (**Table 2**) which could be related to gestation-dependent increase in metabolic enzyme activities and increased blood flow to the liver^55^. Lastly, the cumulative mean fetal AUC for the oral formulations within a 28-day period was lower for both drugs when compared to the therapeutic-equivalent LAI formulation for a similar duration.

Zheng, et al. ^68^ reported a full-body PBPK model to investigate changes in systemic exposure of oral olanzapine during pregnancy. Their predicted plasma drug concentrations did not reach the therapeutic level of 20 ng/mL required for effective treatment after the administration of 10 mg oral olanzapine. Though the predicted C_max_ achieved with oral 10 mg olanzapine in our study was well above the effective therapeutic level, overall drug plasma concentration was lower than what is needed for effective treatment in both second and third trimester. In contrast, the whole-body PBPK model utilized by Zheng, et al. ^50^ predicted a gradual decline in maternal plasma drug concentrations as pregnancy progresses, a trend that was also observed in our predictions for aripiprazole. While both studies did not examine fetal exposures, our study investigated and compared extent of fetal exposure between the oral and LAI formulations.

Though there has been no association between the use of oral and LAI aripiprazole with incidence of congenital malformations^70,71^, there are numerous clinical studies linking olanzapine with increased risk of musculoskeletal defects^72^. While *in utero* fetal exposure to maternal medication does not necessarily translate to risk. Direct effect on the placenta has been associated with the risk associated with some drugs. Hence, data on fetal exposure should be interpreted in a broader context that includes possible effect on the placenta and benefit-risk assessments.

The findings reported here need to be interpreted in the context of certain limitations. Contributions of placenta drug metabolism and transport which may affect the overall fetal drug exposure were not accounted for. From available literature, both aripiprazole ^73^ and olanzapine ^74^ are substrates of P-gp (efflux) transporters. However, insufficient data for proper characterisation of their roles. Additionally, the current model did not capture the dehydrogenation component of aripiprazole metabolic pathway which produces dehydro-aripiprazole, a major metabolite that contributes to its pharmacological activity.

Model predictions for olanzapine and aripiprazole suggest that fetal exposure to these LAI antipsychotics is higher compared to their therapeutic-equivalent oral doses. Further research is needed to understand potential differences in the pharmacokinetic and safety profiles of oral formulations compared to long-acting during pregnancy.

## Data availability

The datasets generated and/or analysed during the current study are available from the corresponding author upon reasonable request.

## Code availability

Related model file developed for this study can be obtained from the corresponding author upon reasonable request.

## Acknowledgements

A. Olagunju reports research funding from the Wellcome Trust, including 227288/Z/23/Z (2024-2031).

## Author contributions

PB, MM, SA and AO contributed to the conception and design of this study. PB and MM conducted the study. Data analysis was conducted by PB and MM. The first draft was written by PB, MM and SA, and revised by SA and AO. All authors read and approved the final draft of the manuscript.

## Supplementary Files

**Table S1**: Drug parameters inputted in the model.

**Table S2:** Validation of whole-body adult PBPK model predicted PK parameters of oral and LAI aripiprazole and olanzapine to observed data.

**Table S3:** Predicted vs observed plasma concentration of oral aripiprazole and olanzapine in the pregnant population in each trimester.

**Table S4:** Predicted vs observed PK parameters for oral aripiprazole and olanzapine at delivery in mother and fetus.

**Figure S1**: Predicted average cord-to-maternal plasma drug concentration ratios of oral (PO) and their therapeutic-equivalent long-acting injectable (LAI) doses of aripiprazole during pregnancy in one dosing interval at steady-state.

**Figure S2**: Predicted average cord-to-maternal plasma drug concentration ratios of (PO) and their therapeutic-equivalent long-acting injectable (LAI) doses of olanzapine in pregnancy in one dosing interval at steady-state.

## References

1 Calabrese, J. & Al Khalili, Y. in StatPearls (StatPearls Publishing Copyright © 2024, StatPearls Publishing LLC., 2024).

2 Leucht, S. & Heres, S. Epidemiology, clinical consequences, and psychosocial treatment of nonadherence in schizophrenia. J Clin Psychiatry 67 Suppl 5, 3–8 (2006).

3 Palmer, B. A., Pankratz, V. S. & Bostwick, J. M. The Lifetime Risk of Suicide in Schizophrenia: A Reexamination. Archives of General Psychiatry 62, 247–253 (2005). 10.1001/archpsyc.62.3.247

4 Lawrence, D. & Kisely, S. Review: Inequalities in healthcare provision for people with severe mental illness. Journal of Psychopharmacology 24, 61–68 (2010). 10.1177/1359786810382058

5 Corrigan, P. W. & Watson, A. C. Understanding the impact of stigma on people with mental illness. World Psychiatry 1, 16–20 (2002).

6 Perlick, D. A. et al. Caregiver burden as a predictor of depression among family and friends who provide care for persons with bipolar disorder. Bipolar Disord 18, 183–191 (2016). 10.1111/bdi.12379

7 Yee, N. et al. A meta-analysis of the relationship between psychosis and any type of criminal offending, in both men and women. Schizophrenia Research 220, 16–24 (2020).10.1016/j.schres.2020.04.009

8 Semahegn, A. et al. Psychotropic medication non-adherence and its associated factors among patients with major psychiatric disorders: a systematic review and meta-analysis. Systematic reviews 9, 1–18 (2020).

9 Haddad, P. M., Brain, C. & Scott, J. Nonadherence with antipsychotic medication in schizophrenia: challenges and management strategies. Patient related outcome measures, 43–62 (2014).

10 Mohammed, F., Geda, B., Yadeta, T. A. & Dessie, Y. Antipsychotic medication nonadherence and factors associated among patients with schizophrenia in eastern Ethiopia. BMC psychiatry 24, 108 (2024).

11 Chordiya, M., Gangurde, H. & Borkar, V. Technologies, optimization and analytical parameters in gastroretentive drug delivery systems. Current science, 946–953 (2017).

12 Doherty, M. M. & Pang, K. S. First-pass effect: significance of the intestine for absorption and metabolism. Drug Chem Toxicol 20, 329–344 (1997). 10.3109/01480549709003891

13 Tam, Y. K. Individual variation in first-pass metabolism. Clin Pharmacokinet 25, 300–328 (1993). 10.2165/00003088-199325040-00005

14 Pond, S. M. & Tozer, T. N. First-pass elimination. Basic concepts and clinical consequences. Clin Pharmacokinet 9, 1–25 (1984). 10.2165/00003088-198409010-00001

15 Thompson, C. The use of high-dose antipsychotic medication. The British Journal of Psychiatry 164, 448–458 (1994).

16 Davis, J. M. & Chen, N. Dose response and dose equivalence of antipsychotics. Journal of clinical psychopharmacology 24, 192–208 (2004).

17 McEvoy, J. P. Risks versus benefits of different types of long-acting injectable antipsychotics. J Clin Psychiatry 67 Suppl 5, 15–18 (2006).

18 Gerlach, J. Depot neuroleptics in relapse prevention: advantages and disadvantages. Int Clin Psychopharmacol 9 Suppl 5, 17–20 (1995). 10.1097/00004850-199501005-00004

19 Remington, G. J. & Adams, M. E. Depot neuroleptic therapy: clinical considerations. Can J Psychiatry 40, S5–11 (1995).

20 Brissos, S., Veguilla, M. R., Taylor, D. & Balanzá-Martinez, V. The role of long-acting injectable antipsychotics in schizophrenia: a critical appraisal. Ther Adv Psychopharmacol 4, 198–219 (2014). 10.1177/2045125314540297

21 Dencker, S. J. The risk/benefit ratio of depot neuroleptics: a Scandinavian perspective. J Clin Psychiatry 45, 22–27 (1984).

22 Marder, S. R., Hubbard, J. W., Van Putten, T. & Midha, K. K. Pharmacokinetics of long-acting injectable neuroleptic drugs: clinical implications. Psychopharmacology (Berl) 98, 433–439 (1989). 10.1007/bf00441937

23 Waddell, L. & Taylor, M. Attitudes of patients and mental health staff to antipsychotic long-acting injections: systematic review. Br J Psychiatry Suppl 52, S43–50 (2009). 10.1192/bjp.195.52.s43

24 Olfson, M., Mechanic, D., Hansell, S., Boyer, C. A. & Walkup, J. Prediction of homelessness within three months of discharge among inpatients with schizophrenia. Psychiatr Serv 50, 667–673 (1999). 10.1176/ps.50.5.667

25 Cohen, L. S. et al. Relapse of major depression during pregnancy in women who maintain or discontinue antidepressant treatment. Jama 295, 499–507 (2006).

26 Costantine, M. M. Physiologic and pharmacokinetic changes in pregnancy. Front Pharmacol 5, 65 (2014). 10.3389/fphar.2014.00065

27 Pinheiro, E. A. & Stika, C. S. Drugs in pregnancy: Pharmacologic and physiologic changes that affect clinical care. Semin Perinatol 44, 151221 (2020). 10.1016/j.semperi.2020.151221

28 Bjelica, A., Cetkovic, N., Trninic-Pjevic, A. & Mladenovic-Segedi, L. The phenomenon of pregnancy—A psychological view. Ginekologia polska 89, 102–106 (2018).

29 Rahmawati, A. & Wulandari, R. C. L. Influence of Physical and Psychological of Pregnant Women Toward Health Status of Mother and Baby. Jurnal Kebidanan 9, 148–152 (2019).

30 Wesseloo, R. et al. Risk of postpartum relapse in bipolar disorder and postpartum psychosis: a systematic review and meta-analysis. American Journal of Psychiatry 173, 117–127 (2016).

31 Kendig, S. et al. Consensus bundle on maternal mental health: perinatal depression and anxiety. Journal of Obstetric, Gynecologic & Neonatal Nursing 46, 272–281 (2017).

32 Leight, K. L., Fitelson, E. M., Weston, C. A. & Wisner, K. L. Childbirth and mental disorders. International Review of Psychiatry 22, 453–471 (2010).

33 Schrager, N. L. et al. Trends in first-trimester nausea and vomiting of pregnancy and use of select treatments: Findings from the National Birth Defects Prevention Study. Paediatric and perinatal epidemiology 35, 57–64 (2021).

34 Einarson, A. & Boskovic, R. Use and safety of antipsychotic drugs during pregnancy. Journal of Psychiatric Practice® 15, 183–192 (2009).

35 Galbally, M., Snellen, M. & Power, J. Antipsychotic drugs in pregnancy: a review of their maternal and fetal effects. Therapeutic advances in drug safety 5, 100–109 (2014).

36 Edinoff, A. N. et al. Antipsychotic Use in Pregnancy: Patient Mental Health Challenges, Teratogenicity, Pregnancy Complications, and Postnatal Risks. Neurol Int 14, 62–74 (2022). 10.3390/neurolint14010005

37 Dallmann, A., Ince, I., Coboeken, K., Eissing, T. & Hempel, G. A physiologically based pharmacokinetic model for pregnant women to predict the pharmacokinetics of drugs metabolized via several enzymatic pathways. Clinical pharmacokinetics 57, 749–768 (2018).

38 Zhang, Z. et al. Development of a novel maternal-fetal physiologically based pharmacokinetic model I: insights into factors that determine fetal drug exposure through simulations and sensitivity analyses. Drug Metabolism and Disposition 45, 920–938 (2017).

39 Atoyebi, S. et al. Physiologically-based pharmacokinetic modelling of long-acting injectable cabotegravir and rilpivirine in pregnancy. British Journal of Clinical Pharmacology (2024).

40 Otsuka. Abilify Maintena® (aripiprazole). Prescribing Information., <https://www.otsuka-us.com/sites/g/files/qhldwo9046/files/media/static/Abilify-PI.pdf> (2002).

41 Aristada TM. Aristada TM (aripiprazole lauroxil). Prescribing Information. Alkermes, Inc., <https://www.aristada.com/downloadables/ARISTADA-PI.pdf> (2001).

42 Raoufinia, A. et al. Aripiprazole once-monthly 400 mg: comparison of pharmacokinetics, tolerability, and safety of deltoid versus gluteal administration. International Journal of Neuropsychopharmacology 20, 295–304 (2017).

43 Mallikaarjun, S. et al. Pharmacokinetics, tolerability and safety of aripiprazole once-monthly in adult schizophrenia: an open-label, parallel-arm, multiple-dose study. Schizophrenia research 150, 281–288 (2013).

44 Samalin, L., Garay, R., Ameg, A. & Llorca, P.-M. Olanzapine pamoate for the treatment of schizophrenia – a safety evaluation. Expert Opinion on Drug Safety 15, 403–411 (2016). 10.1517/14740338.2016.1141893

45 Birnbaum L et al. Physiological parameter values for PBPK models. A report prepared by the International Life Sciences Institute Risk Science Institute.. (Washington, DC, 1994).

46 Lawrence, X. Y. & Amidon, G. L. A compartmental absorption and transit model for estimating oral drug absorption. International journal of pharmaceutics 186, 119–125 (1999).

47 Gertz, M., Harrison, A., Houston, J. B. & Galetin, A. Prediction of human intestinal first-pass metabolism of 25 CYP3A substrates from in vitro clearance and permeability data. Drug Metabolism and Disposition 38, 1147–1158 (2010).

48 Sun, L., von Moltke, L. & Rowland Yeo, K. Application of Physiologically Based Pharmacokinetic Modeling to Predict the Effect of Renal Impairment on the Pharmacokinetics of Olanzapine and Samidorphan Given in Combination. Clinical Pharmacokinetics 60, 637–647 (2021). 10.1007/s40262-020-00969-w

49 Paine, M. F. et al. The human intestinal cytochrome P450 “pie”. Drug metabolism and disposition 34, 880–886 (2006).

50 Zheng, L. et al. Dose adjustment of quetiapine and aripiprazole for pregnant women using physiologically based pharmacokinetic modeling and simulation. Clinical Pharmacokinetics 60, 623–635 (2021).

51 Poulin, P. & Theil, F. P. Prediction of Pharmacokinetics Prior to In Vivo Studies. 1. Mechanism-Based Prediction of Volume of Distribution. Journal of Pharmaceutical Sciences 91, 129–156 (2002). 10.1002/jps.10005

52 Atoyebi, S. A. et al. Using mechanistic physiologically-based pharmacokinetic models to assess prenatal drug exposure: Thalidomide versus efavirenz as case studies. Eur J Pharm Sci 140, 105068 (2019). 10.1016/j.ejps.2019.105068

53 Dallmann, A. et al. Physiologically Based Pharmacokinetic Modeling of Renally Cleared Drugs in Pregnant Women. Clin Pharmacokinet 56, 1525–1541 (2017). 10.1007/s40262-017-0538-0

54 FDA. HIGHLIGHTS OF PRESCRIBING INFORMATION FOR ABILIFY®, <https://www.accessdata.fda.gov/drugsatfda_docs/label/2014/021436s038,021713s030,021729s022,021866s023lbl.pdf> (2002).

55 Abduljalil, K., Furness, P., Johnson, T. N., Rostami-Hodjegan, A. & Soltani, H. Anatomical, physiological and metabolic changes with gestational age during normal pregnancy: a database for parameters required in physiologically based pharmacokinetic modelling. Clinical pharmacokinetics 51, 365–396 (2012).

56 Valentin, J. Basic anatomical and physiological data for use in radiological protection: reference values: ICRP Publication 89. Annals of the ICRP 32, 1–277 (2002).

57 Abduljalil, K., Cain, T., Humphries, H. & Rostami-Hodjegan, A. Deciding on Success Criteria for Predictability of Pharmacokinetic Parameters from In Vitro Studies: An Analysis Based on In Vivo Observations. Drug Metabolism and Disposition 42, 1478–1484 (2014). 10.1124/dmd.114.058099

58 Boulton, D. W. et al. Pharmacokinetics and tolerability of intramuscular, oral and intravenous aripiprazole in healthy subjects and in patients with schizophrenia. Clinical pharmacokinetics 47, 475–485 (2008).

59 Wojnicz, A. et al. Effective phospholipids removing microelution-solid phase extraction LC-MS/MS method for simultaneous plasma quantification of aripiprazole and dehydro-aripiprazole: Application to human pharmacokinetic studies. Journal of Pharmaceutical and Biomedical Analysis 151, 116–125 (2018).

60 Westin, A. A. et al. Treatment With Antipsychotics in Pregnancy: Changes in Drug Disposition. Clin Pharmacol Ther 103, 477–484 (2018). 10.1002/cpt.770

61 Griffiths, S. K. & Campbell, J. P. Placental structure, function and drug transfer. Continuing Education in Anaesthesia, Critical Care & Pain 15, 84–89 (2015).

62 Hines, R. N. Ontogeny of human hepatic cytochromes P450. Journal of biochemical and molecular toxicology 21, 169–175 (2007).

63 Barter, Z. E. et al. Covariation of human microsomal protein per gram of liver with age: absence of influence of operator and sample storage may justify interlaboratory data pooling. Drug Metabolism and Disposition 36, 2405–2409 (2008).

64 Icrp, A. Basic anatomical and physiological data for use in radiological protection: reference values. Ann. ICRP 32, 1–277 (2002).

65 Aichhorn, W. et al. Olanzapine plasma concentration in a newborn. J Psychopharmacol 22, 923–924 (2008). 10.1177/0269881107083849

66 Detke, H. C., Zhao, F., Garhyan, P., Carlson, J. & McDonnell, D. Dose correspondence between olanzapine long-acting injection and oral olanzapine: recommendations for switching. International Clinical Psychopharmacology 26, 35–42 (2011). 10.1097/YIC.0b013e32834093d1

67 Miron, A.-A. et al. Switch from Olanzapine Long-Acting Injectable to its Oral Equivalent during COVID-19 Pandemic: a Real World Observational Study. Psychiatric Quarterly 93, 627–635 (2022). 10.1007/s11126-021-09924-9

68 Zheng, L. et al. Physiologically Based Pharmacokinetic Modeling in Pregnant Women Suggests Minor Decrease in Maternal Exposure to Olanzapine. Front Pharmacol 12, 793346 (2021). 10.3389/fphar.2021.793346

69 Correll, C. U. et al. Pharmacokinetic Characteristics of Long-Acting Injectable Antipsychotics for Schizophrenia: An Overview. CNS Drugs 35, 39–59 (2021). 10.1007/s40263-020-00779-5

70 Eleftheriou, G., Butera, R., Sangiovanni, A., Palumbo, C. & Bondi, E. Long-Acting Injectable Antipsychotic Treatment during Pregnancy: A Case Series. Int J Environ Res Public Health 20 (2023). 10.3390/ijerph20043080

71 Bellet, F. et al. Exposure to aripiprazole during embryogenesis: a prospective multicenter cohort study. Pharmacoepidemiology and Drug Safety 24, 368–380 (2015).

72 Ellfolk, M. et al. Second-generation antipsychotic use during pregnancy and risk of congenital malformations. European Journal of Clinical Pharmacology 77, 1737–1745 (2021). 10.1007/s00228-021-03169-y

73 Wang, J. S. et al. Aripiprazole brain concentration is altered in P-glycoprotein deficient mice. Schizophr Res 110, 90–94 (2009). 10.1016/j.schres.2009.01.011

74 Xu, Y. et al. P-glycoprotein mediates the pharmacokinetic interaction of olanzapine with fluoxetine in rats. Toxicology and Applied Pharmacology 431, 115735 (2021). 10.1016/j.taap.2021.115735

75 Wishart, D. S. et al. DrugBank 5.0: a major update to the DrugBank database for 2018. Nucleic acids research 46, D1074–D1082 (2018).

76 Sun, L., von Moltke, L. & Rowland Yeo, K. Physiologically-Based Pharmacokinetic Modeling for Predicting Drug Interactions of a Combination of Olanzapine and Samidorphan. CPT: Pharmacometrics & Systems Pharmacology 9, 106–114 (2020). 10.1002/psp4.12488

77 Kneller, L. A., Zubiaur, P., Koller, D., Abad-Santos, F. & Hempel, G. Influence of CYP2D6 Phenotypes on the Pharmacokinetics of Aripiprazole and Dehydro-Aripiprazole Using a Physiologically Based Pharmacokinetic Approach. Clin Pharmacokinet 60, 1569–1582 (2021). 10.1007/s40262-021-01041-x

78 Mallikaarjun, S., Shoaf, S. E., Boulton, D. W. & Bramer, S. L. Effects of hepatic or renal impairment on the pharmacokinetics of aripiprazole. Clinical pharmacokinetics 47, 533–542 (2008).

79 Korprasertthaworn, P. et al. In Vitro Characterization of the Human Liver Microsomal Kinetics and Reaction Phenotyping of Olanzapine Metabolism. Drug Metab Dispos 43, 1806–1814 (2015). 10.1124/dmd.115.064790

80 Vieira, M. d. L. et al. PBPK model describes the effects of comedication and genetic polymorphism on systemic exposure of drugs that undergo multiple clearance pathways. Clinical Pharmacology & Therapeutics 95, 550–557 (2014).

81 Wang, X. et al. Population pharmacokinetic modeling and exposure-response analysis for aripiprazole once monthly in subjects with schizophrenia. Clinical Pharmacology in Drug Development 11, 150–164 (2022).

82 Heres, S., Kraemer, S., Bergstrom, R. F. & Detke, H. C. Pharmacokinetics of olanzapine long-acting injection: the clinical perspective. Int Clin Psychopharmacol 29, 299–312 (2014). 10.1097/yic.0000000000000040

83 Du, P. et al. Open-Label, Randomized, Single-Dose, 2-Period, 2-Sequence Crossover, Comparative Pharmacokinetic Study to Evaluate Bioequivalence of 2 Oral Formulations of Olanzapine Under Fasting and Fed Conditions. Clinical Pharmacology in Drug Development 9, 621–628 (2020). 10.1002/cpdd.743

84 Sun, L., McDonnell, D., Liu, J. & von Moltke, L. Bioequivalence of Olanzapine Given in Combination With Samidorphan as a Bilayer Tablet (ALKS 3831) Compared With Olanzapine-Alone Tablets: Results From a Randomized, Crossover Relative Bioavailability Study. Clinical Pharmacology in Drug Development 8, 459–466 (2019). 10.1002/cpdd.601

85 Mitchell, M. et al. Single- and Multiple-Dose Pharmacokinetic, Safety, and Tolerability Profiles of Olanzapine Long-Acting Injection: An Open-Label, Multicenter, Nonrandomized Study in Patients With Schizophrenia. Clinical Therapeutics 35, 1890–1908 (2013). 10.1016/j.clinthera.2013.09.023

86 Westin, A. A. et al. Treatment with antipsychotics in pregnancy: changes in drug disposition. Clinical Pharmacology & Therapeutics 103, 477–484 (2018).

87 Nguyen, T., Teoh, S., Hackett, L. P. & Ilett, K. Placental transfer of aripiprazole. Australian and New Zealand Journal of Psychiatry 45, 500–501 (2011).

