## Supplemental Tables and Figures for "Comparative modeling of fetal exposure to maternal long-acting injectable versus oral daily antipsychotics"

**Supplementary information**

Table S1: Drug parameters inputted in the model.

| **Drug Parameter** | **Aripiprazole** | **Olanzapine** |
|---|---|---|
| $pKa$ | 7.46 (1) | 7.24 (2) |
| $f_{up}$ | 0.01 (1) | 0.07 (2) |
| $Caco-2$ (cm/sec) | 0.000047 (3) | - |
| $LogP_{o:w}$ | 5.11 (4) | 2.89 (2) |
| $PSA$ (Å2) | 44.81 (1) | - |
| $HBD$ | 1 (1) | - |
| $R$ | 0.61 (3) | 0.62 (2) 0.62(2)0.62(2)0.62 |
| $Cl_{ren}$ (L/h) | 0.003 (5) | - |
| $Cl_{int}CYP3A4$ (L/h) | 8.4 (3) | 4.42 (6)* |
| $Cl_{int}CYP2D6$ (L/h) | 69 (3) | - |
| $Cl_{int}CYP1A2$ (μL/min/mg) | - | 23.67 (6) |
| $Cl_{int}CYP2C8$ (μL/min/mg) | - | 4.98 (6) |
| $Cl_{int}FMO3$ (μL/min/mg) | - | 4.28 (6) |
| $Cl_{int}UGT1A4$ (μL/min/mg) | - | 20.58 (6) |
| $f_mCYP3A4$ | 0.56 (7) | - |
| $f_mCYP2D6$ | 0.43 (7) | - |
| $K_A$ | - | 0.6 (8) |
| $K_{im1}$ | 0.000904 (9) | 9.36e-4 (10) |
| $K_{im2}$ | - | 3.0e-4 (fitted) |
| $CF\_Vd$ | 0.4 (fitted) | 1 (fitted) |

pKa is the acid dissociation constant; $f_{up}$ is the fraction of unbound drug in plasma; Caco-2 is the apparent permeability coefficient measured in Caco-2 cell monolayers; $LogP_{o:w}$ is the logarithm of the partition coefficient between octanol and water, indicating lipophilicity; PSA is the polar surface area of the drug; HBD is the number of hydrogen bond donors in the molecule; R is the blood-to-plasma concentration ratio; $Cl_{ren}$ is the renal clearance rate; $Cl_{int}$CYP3A4 is the intrinsic clearance by the CYP3A4 enzyme; $Cl_{int}$CYP2D6 is the intrinsic clearance by the CYP2D6 enzyme; $Cl_{int}$CYP1A2 is the intrinsic clearance by the CYP1A2 enzyme; $Cl_{int}$CYP2C8 is the intrinsic clearance by the CYP2C8 enzyme; $Cl_{int}$FMO3 is the intrinsic clearance by the FMO3 enzyme; $Cl_{int}$UGT1A4 is the intrinsic clearance by the UGT1A4 enzyme; $f_m$CYP3A4 is the fraction metabolized by CYP3A4, $f_m$CYP2D6 is the fraction metabolized by CYP2D6; $K_{im}$ is the first-order release rate constant for the intramuscular depot and CF_Vd is the correction factor for the volume of distribution.

*Unit for parameter is µL/min/mg

**Table S2:** Validation of whole-body adult PBPK model predicted PK parameters of oral and LAI aripiprazole and olanzapine to observed data.

| Pharmacokinetic Parameter | Observed | Simulated | AAFE | References |
|---|---|---|---|---|
| **Oral doses** | | | | |
| **5 mg ARI oral single dose** | **(n = 14)** | **(n = 100)** | | (11) |
| $AUC_{0\text{-}384h}$ (ng.h/mL) | 1108 | 1042 | 1.06 | |
| $C_{max}$ (ng/mL) | 19.9 | 11 | 1.81 | |
| $T_{1/2}$ (h) | 103.4 | 58.7 | 1.76 | |
| **10 mg ARI oral single dose under fasting conditions** | **(n = 103)** | **(n = 100)** | | (12) |
| $AUC_{0\text{-}72h}$ (ng.h/mL) | 1944 | 1385 | 1.40 | |
| $C_{max}$ (ng/mL) | 55.7 | 28.9 | 1.93 | |
| $T_{1/2}$ (h) | 53.2 | 60.1 | 1.13 | |
| **5 mg OLZ oral daily under fasting conditions** | **(n = 28)** | **(n =100)** | | (13) |
| $AUC_{0\text{-}144h}$ (ng.h/mL) | 317 | 266 | 1.19 | |
| $C_{max}$ (ng/mL) | 7.8 | 13.5 | 1.73 | |
| CL (L/h) | 16.7 | 23.3 | 1.40 | |
| **5 mg OLZ oral daily under fed conditions** | **(n = 21)** | **(n = 100)** | | (13) |
| $AUC_{0\text{-}144}$ (ng.h/mL) | 275 | 273 | 1.16 | |
| $C_{max}$ (ng/mL) | 6.6 | 11.6 | 1.49 | |
| CL (L/h) | 15.8 | 22.8 | 1.44 | |
| **10 mg ARI oral daily dose** | **(n = 45)** | **(n = 100)** | | (14) |
| $AUC_{0\text{-}168}$ (ng.h/mL) | 599 | 536 | 1.12 | |
| $C_{max}$ (ng/mL) | 16.7 | 17.9 | 1.07 | |
| CL/F (L/h) | 18.2 | 18.7 | 1.03 | |
| **Long-acting injectable doses** | | | | |
| **400 mg ARI gluteal IM single dose** | **(n = 19)** | **(n = 100)** | | (15) |
| $AUC_{0\text{-}126d}$ (ng.day/mL) | 7340 | 8828 | 1.20 | |
| $AUC_{0\text{-}28d}$ (ng.d/mL) | 2380 | 3756 | 1.58 | |
| $C_{max}$ (ng/mL) | 136 | 164 | 1.20 | |
| $C_{28d}$ (ng/mL) | 86.2 | 124 | 1.44 | |
| $T_{1/2}$ (days) | 24 | 32 | 1.33 | |
| **400 mg ARI deltoid IM single dose** | **(n = 18)** | **(n = 100)** | | (15) |
| $AUC_{0\text{-}126d}$ (ng.day/mL) | 7360 | 8828 | 1.20 | |

| | | | | |
|---|---|---|---|---|
| $AUC_{0\text{-}28d}$ (ng.day/mL) | 3120 | 3756 | 1.20 | |
| $C_{max}$ (ng/mL) | 170 | 164 | 1.04 | |
| $C_{28d}$ (ng/mL) | 103 | 124 | 1.20 | |
| $T_{1/2}$ (days) | 17.8 | 32.0 | 1.80 | |
| **400 mg ARI gluteal/deltoid IM single once-monthly LAI dose[a]** | **(n = 68)** | **(n = 100)** | | (15) |
| $AUC_{0\text{-}28d}$ (ng.day/mL) | 2359 | 3757 | 1.59 | |
| $C_{max}$ (ng/mL) | 126 | 169 | 1.34 | |
| $C_{28d}$ (ng/mL) | 79 | 125 | 1.58 | |
| **400 mg ARI deltoid/deltoid IM single once-monthly LAI dose[a]** | **(n = 73)** | **(n = 100)** | | (15) |
| $AUC_{0\text{-}28d}$ (ng.day/mL) | 2728 | 3757 | 1.38 | |
| $C_{max}$ (ng/mL) | 135 | 169 | 1.25 | |
| $C_{28d}$ (ng/mL) | 112 | 125 | 1.11 | |
| **400 mg ARI combined gluteal/deltoid and deltoid/deltoid IM once-monthly multiple doses after fifth injection[b]** | **-** | **(n = 100)** | | (15) |
| $AUC_{112\text{-}140d}$ (ng.day/mL) | 7027[c] | 8916 | 1.27 | |
| $C_{max,ss}$ (ng/mL) | 328[d] | 376 | 1.15 | |
| $C_{min,ss}$ (ng/mL) | 239[e] | 247 | 1.03 | |
| **300 mg OLZ bi-weekly IM dose at steady-state** | **(n = 19)** | **(n = 100)** | | (16) |
| $AUC_{T,SS}$ (ng·h/mL) | 12400 | 10083 | 1.23 | |
| $C_{max,ss}$ (ng/mL) | 46.1 | 74.8 | 1.62 | |
| $C_{mean,ss}$ (ng/mL) | 35.2 | 35.9 | 1.02 | |
| **405 mg OLZ bi-weekly IM dose at steady-state** | **(n = 29)** | **(n = 100)** | | (16) |
| $AUC_{T,\,SS}$ (ng·h/mL) | 23600 | 20797 | 1.14 | |
| $C_{max,ss}$ (ng/mL) | 47.6 | 63.5 | 1.33 | |
| $C_{mean,ss}$ (ng/mL) | 37 | 42 | 1.14 | |
| $CL_{ss}/F$ (L/h) | 17.1 | 19.4 | 1.14 | |

Data are presented as means. AUC: area under the drug plasma concentration curve, $C_{max}$: maximum plasma concentration, $C_{max,ss}$: maximum plasma concentration at steady state, $C_{min,ss}$: minimum plasma concentration at steady state, $C_{28}$: plasma concentration at 28 days post-dose, IM: intramuscular, LAI: long-acting injectable, $AUC_{T,SS}$: area under the curve at steady state during one dosing interval; $C_{max,ss}$: maximum concentration at steady-state; $C_{mean,SS}$: mean olanzapine concentration at steady state; CL: clearance; CLss/F: apparent clearance at steady-state, AAFE: absolute average fold error, ARI: aripiprazole, OLZ: olanzapine.

[a]Patients were randomised to receive their first injection of aripiprazole 400 mg in either the

deltoid or gluteal muscle, followed by four monthly injections in the deltoid.
[b]Both cohorts receiving a deltoid or gluteal first injection followed by four monthly deltoid injections were combined for analysis.
[c]n = 36
[d]n = 39
[e]n = 86

**Table S3:** Predicted vs observed plasma concentration of oral aripiprazole and olanzapine in the pregnant population in each trimester.

| Pharmacokinetic parameter | Observed | Simulated | AAFE | References |
|---|---|---|---|---|
| **15 mg ARI oral daily dose** | **(n = 14)** | **(n = 100)** | | (17) |
| 1st trimester (ng/mL) | 153.02[a] | 82.82 | 1.85 | |
| 2nd trimester (ng/mL) | 108.76[a] | 63.16 | 1.72 | |
| 3rd trimester (ng/mL) | 76.79[a] | 48.85 | 1.57 | |
| **10 mg OLZ oral daily dose** | **(n = 29)** | **(n = 100)** | | (17) |
| 1st trimester (ng/mL) | 20.9 | 19.1 | 1.09 | |
| 2nd trimester (ng/mL) | 20.1 | 17.1 | 1.18 | |
| 3rd trimester (ng/mL) | 19.3 | 15.5 | 1.25 | |

Data are presented as means. ARI: aripiprazole, OLZ: olanzapine, AAFE: absolute average fold error.

[a]Values were calculated as fractions of reported concentrations of the active moiety of aripiprazole in the plasma.

**Table S4:** Predicted vs observed PK parameters for oral aripiprazole and olanzapine at delivery in mother and fetus.

| Pharmacokinetic Parameter | Observed | Simulated | AAFE | References |
|---|---|---|---|---|
| **10 mg ARI oral daily dose** | **(37 – 39.3 wks)** | **(37– 40 wks)** | | (18) |
| Cord plasma concentration (µg/L) | 45[a] | 27.5[b] | 1.64 | |
| Maternal plasma concentration (µg/L) | 70[a] | 40.9[b] | 1.71 | |
| Cord : maternal plasma drug ratio | 0.64[a] | 0.67[b] | 1.047 | |
| **10 mg OLZ oral daily dose** | **(n = 1)** | **(n = 100)** | | (19) |
| Fetal plasma concentration (ng/mL) | 35 | 23.6 | 1.48 | |
| Maternal plasma concentration (ng/mL) | 11 | 9 | 1.22 | |

Data are presented as means. AAFE: absolute average fold error, ARI: aripiprazole, OLZ: olanzapine.

[a]n = 1

[b]n =100

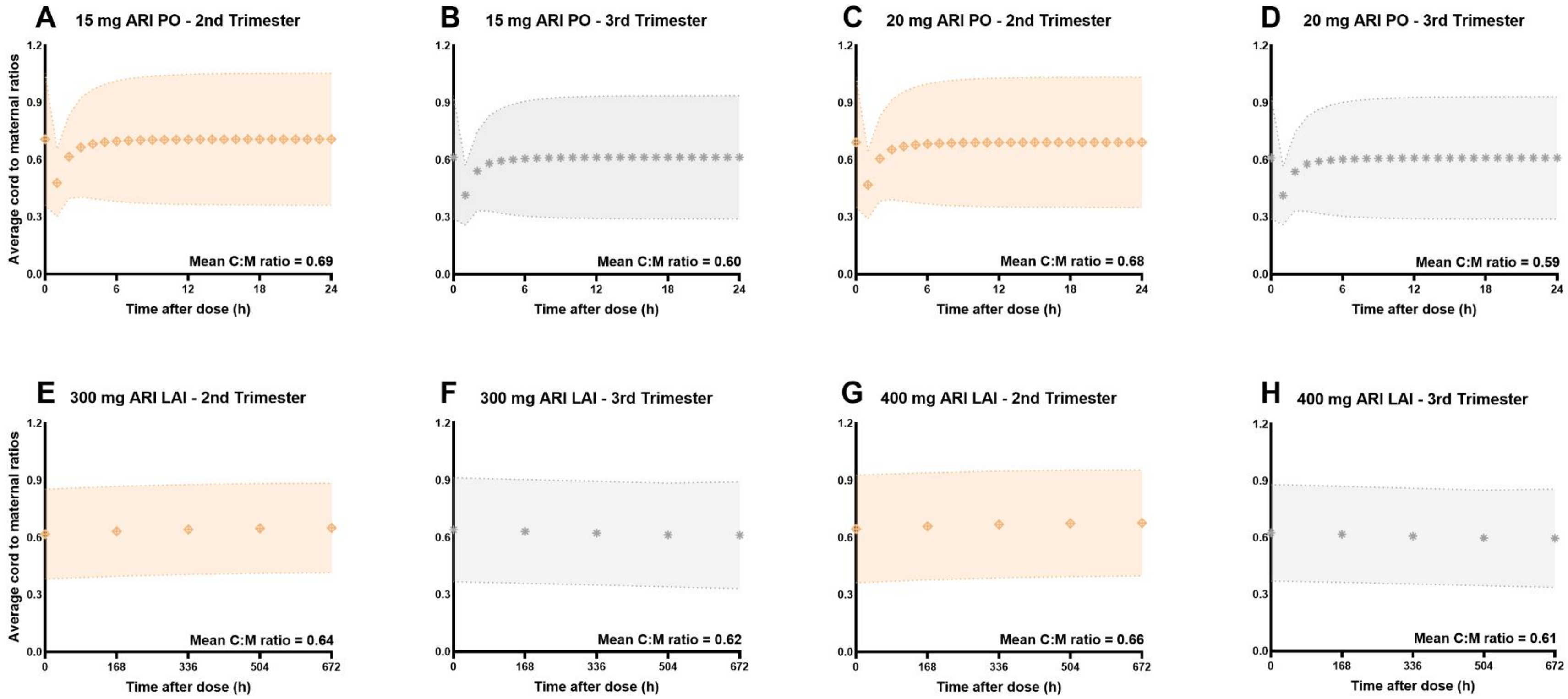


**Figure S1**: Predicted average cord-to-maternal plasma drug concentration ratios of oral (PO) and their therapeutic-equivalent long-acting injectable (LAI) doses of aripiprazole during pregnancy in one dosing interval at steady-state. (A) 15 mg oral dose in the second trimester, (B) 15 mg oral dose in the third trimester, (C) 20 mg oral dose in the second trimester, (D) 20 mg oral dose in the third trimester (E) 300 mg LAI dose in the second trimester, (F) 300 mg LAI dose in the third trimester, , (G) 400 mg LAI dose in the second trimester, (H) 400 mg LAI in the third trimester.

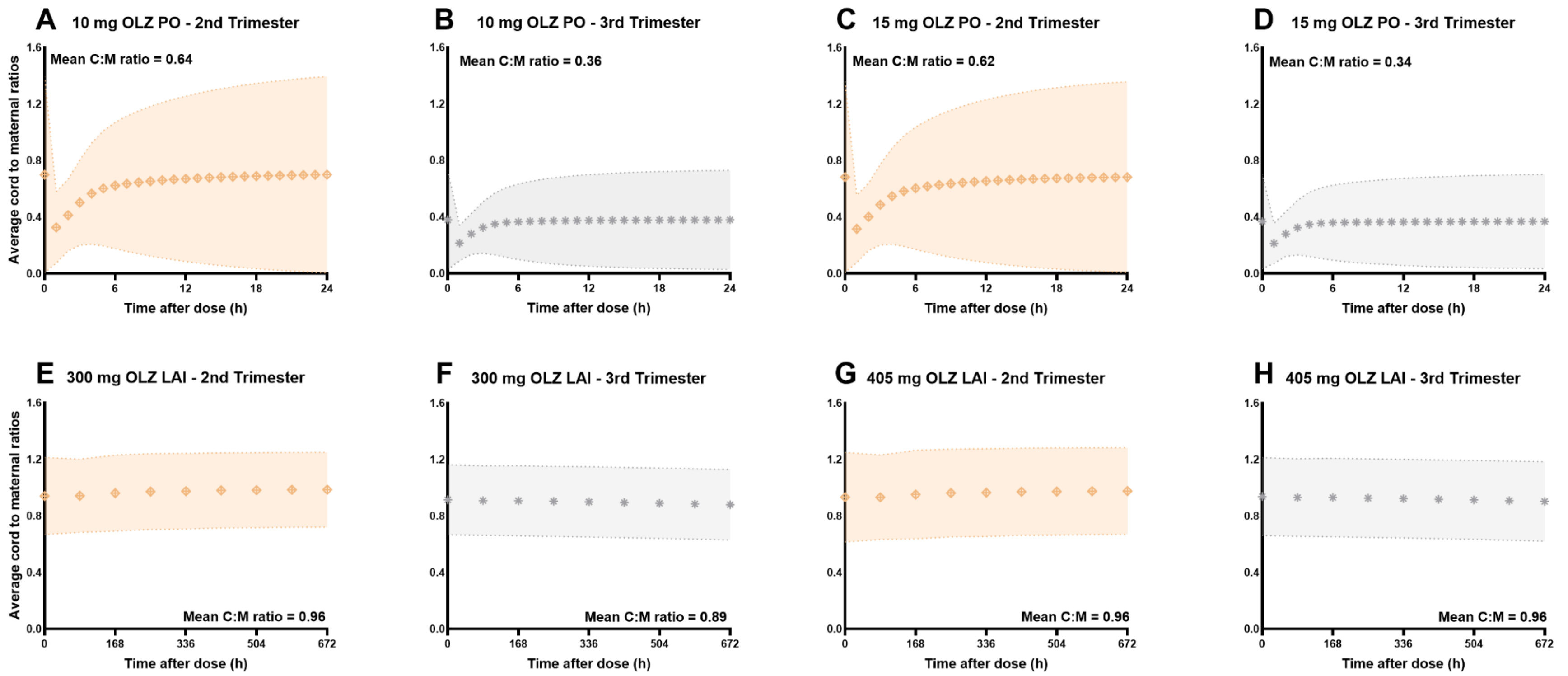


**Figure S2**: Predicted average cord-to-maternal plasma drug concentration ratios of (PO) and their therapeutic-equivalent long-acting injectable (LAI) doses of olanzapine in pregnancy in one dosing interval at steady-state. (A) 10 mg oral dose in the second trimester, (B) 10 mg oral dose in the third trimester, (C) 15 mg oral dose in the second trimester, (D) 15 mg oral dose in the third trimester, (E) 300 mg LAI dose in the second trimester, (F) 300 mg LAI dose in the third trimester, (G) 405 mg LAI dose in the second trimester, (H) 405 mg LAI in the third trimester.

References


1. Wishart DS, Feunang YD, Guo AC, Lo EJ, Marcu A, Grant JR, et al. DrugBank 5.0: a major update to the DrugBank database for 2018. Nucleic acids research. 2018;46(D1):D1074-D82.
2. Sun L, von Moltke L, Rowland Yeo K. Physiologically-Based Pharmacokinetic Modeling for Predicting Drug Interactions of a Combination of Olanzapine and Samidorphan. CPT: Pharmacometrics & Systems Pharmacology. 2020;9(2):106-14.
3. Zheng L, Tang S, Tang R, Xu M, Jiang X, Wang L. Dose adjustment of quetiapine and aripiprazole for pregnant women using physiologically based pharmacokinetic modeling and simulation. Clinical Pharmacokinetics. 2021;60:623-35.
4. Kneller LA, Zubiaur P, Koller D, Abad-Santos F, Hempel G. Influence of CYP2D6 Phenotypes on the Pharmacokinetics of Aripiprazole and Dehydro-Aripiprazole Using a Physiologically Based Pharmacokinetic Approach. Clin Pharmacokinet. 2021;60(12):1569-82.
5. Mallikaarjun S, Shoaf SE, Boulton DW, Bramer SL. Effects of hepatic or renal impairment on the pharmacokinetics of aripiprazole. Clinical pharmacokinetics. 2008;47:533-42.
6. Korprasertthaworn P, Polasek TM, Sorich MJ, McLachlan AJ, Miners JO, Tucker GT, Rowland A. In Vitro Characterization of the Human Liver Microsomal Kinetics and Reaction Phenotyping of Olanzapine Metabolism. Drug Metab Dispos. 2015;43(11):1806-14.
7. Vieira MdL, Kim MJ, Apparaju S, Sinha V, Zineh I, Huang SM, Zhao P. PBPK model describes the effects of comedication and genetic polymorphism on systemic exposure of drugs that undergo multiple clearance pathways. Clinical Pharmacology & Therapeutics. 2014;95(5):550-7.
8. Sun L, von Moltke L, Rowland Yeo K. Application of Physiologically Based Pharmacokinetic Modeling to Predict the Effect of Renal Impairment on the Pharmacokinetics of Olanzapine and Samidorphan Given in Combination. Clinical Pharmacokinetics. 2021;60(5):637-47.
9. Wang X, Raoufinia A, Bihorel S, Passarell J, Mallikaarjun S, Phillips L. Population pharmacokinetic modeling and exposure-response analysis for aripiprazole once monthly in subjects with schizophrenia. Clinical Pharmacology in Drug Development. 2022;11(2):150-64.
10. Heres S, Kraemer S, Bergstrom RF, Detke HC. Pharmacokinetics of olanzapine long-acting injection: the clinical perspective. Int Clin Psychopharmacol. 2014;29(6):299-312.
11. Boulton DW, Kollia G, Mallikaarjun S, Komoroski B, Sharma A, Kovalick LJ, Reeves RA. Pharmacokinetics and tolerability of intramuscular, oral and intravenous aripiprazole in healthy subjects and in patients with schizophrenia. Clinical pharmacokinetics. 2008;47:475-85.
12. Wojnicz A, Belmonte C, Koller D, Ruiz-Nuño A, Román M, Ochoa D, Abad-Santos F. Effective phospholipids removing microelution-solid phase extraction LC-MS/MS method for simultaneous plasma quantification of aripiprazole and dehydro-aripiprazole: Application to human pharmacokinetic studies. Journal of Pharmaceutical and Biomedical Analysis. 2018;151:116-25.

13. Du P, Li P, Liu H, Zhao R, Zhao Z, Yu W, et al. Open-Label, Randomized, Single-Dose, 2-Period, 2-Sequence Crossover, Comparative Pharmacokinetic Study to Evaluate Bioequivalence of 2 Oral Formulations of Olanzapine Under Fasting and Fed Conditions. Clinical Pharmacology in Drug Development. 2020;9(5):621-8.
14. Sun L, McDonnell D, Liu J, von Moltke L. Bioequivalence of Olanzapine Given in Combination With Samidorphan as a Bilayer Tablet (ALKS 3831) Compared With Olanzapine-Alone Tablets: Results From a Randomized, Crossover Relative Bioavailability Study. Clinical Pharmacology in Drug Development. 2019;8(4):459-66.
15. Raoufinia A, Peters-Strickland T, Nylander A-G, Baker RA, Eramo A, Jin N, et al. Aripiprazole once-monthly 400 mg: comparison of pharmacokinetics, tolerability, and safety of deltoid versus gluteal administration. International Journal of Neuropsychopharmacology. 2017;20(4):295-304.
16. Mitchell M, Kothare P, Bergstrom R, Zhao F, Jen KY, Walker D, et al. Single- and Multiple-Dose Pharmacokinetic, Safety, and Tolerability Profiles of Olanzapine Long-Acting Injection: An Open-Label, Multicenter, Nonrandomized Study in Patients With Schizophrenia. Clinical Therapeutics. 2013;35(12):1890-908.
17. Westin AA, Brekke M, Molden E, Skogvoll E, Castberg I, Spigset O. Treatment with antipsychotics in pregnancy: changes in drug disposition. Clinical Pharmacology & Therapeutics. 2018;103(3):477-84.
18. Nguyen T, Teoh S, Hackett LP, Ilett K. Placental transfer of aripiprazole. Australian and New Zealand Journal of Psychiatry. 2011;45(6):500-1.
19. Aichhorn W, Yazdi K, Kralovec K, Steiner H, Whitworth S, Stuppaeck C. Olanzapine plasma concentration in a newborn. J Psychopharmacol. 2008;22(8):923-4.
